# The Lived Experience of Mothers of Children with Cancer

**DOI:** 10.1101/2024.06.26.24309392

**Authors:** Lidya K. Seyoum, Adugnaw Berhane, Eshetu Girma

**Affiliations:** School of Public Health, Addis Ababa University, Ethiopia

**Author notes:** Corresponding Author: Lidya K. Seyoum, School of Public Health, Addis Ababa University, Ethiopia.

**Keywords:** Interpretive phenomenological analysis, childhood cancer, Ethiopia

## Abstract

**Background:** Thus, the path of mothers with cancer children is intricate and severe. Several studies have addressed this topic. Most have considered the long-term effects of the children’s cancer diagnosis on the mother from a mental health perspective. The experience of living with cancer children as mothers reveals the urgent need to pay attention to the mother’s emotional, psychological, and social factors shaping caregiving.

**Objective:** This study aimed to look further into the lived experiences of Ethiopian mothers with children suffering from childhood cancer.

**Method:** Interpretive phenomenological approach of qualitative study conducted at the pediatric oncology unit of Tikur Anbessa Specialized Hospital and Tesfa Addis Parents Childhood Organization. Study participants were selected using the snowball purposive sampling technique. Data were collected using semi-structured interviews. A total of 12 mothers were interviewed. Interpretative phenomenological analysis was employed and Open Code 4.3 software was used for data management and used thematic analysis.

**Result:** When raising a child with childhood cancer, mothers encounter a wide range of issues in the context of childhood cancer care, including fatigue, poor access to high-quality medical care, divorce, parenting withdrawal, excessive medical expenses, lack of sufficient and accurate information, anxiety, impatience, forgetfulness, regret, and spiritual struggles. To overcome such difficulties, mothers employ loans, financial aid, and the sale of their homes and other assets to reduce their financial burden. Mothers’ other coping method was to accept many sorts of support from family, friends, strangers, and organizations. The assistance received helped mothers gain resilience.

**Conclusion:** The study provides crucial insights into the experiences of mothers of children with childhood cancer. The diagnosis of childhood cancer has a profound impact on mothers, and it elicits various reactions from them. Investigate the emotional, mental, and social drivers of caregiving, emphasizing important factors such as financial pressure, insufficient quality medical treatment, emotional disturbances, and strategies of coping with their children by the mothers. Establish counseling and support schemes for mothers to cope with cancer therapy for their child. Organize community awareness campaigns to increase understanding, reduce stigma, and promote early treatment.

## Background

Childhood cancer is a word used to characterize malignancies that develop from birth up to the age of 14 years. Childhood cancers are extremely rare and may differ from adult cancers in their invasion, metastasis, therapeutic interventions, and treatment outcome(1,2). According to the World Health Organisation (WHO) in the year of 2020 GC, 206,362 incidence of childhood cancer were registered worldwide, based on this report Africa is the second big contributor to the number following Asia(3). According to estimates, Ethiopia is the eighth leading contributor to global childhood cancer incidence and the third biggest contributor in Africa, trailing only Nigeria and Egypt, contributing 5,141 new childhood cancer cases in the year(3).The prevalence of childhood cancer in Ethiopia is 2,520 in 2020(3).Given the severity of the condition and the challenges it poses to the family, the figure is concerning.

In Sub-Saharan Africa, the care of children with malignant solid tumors is jeopardized by resource constraints spanning from insufficient healthcare resources and a scarcity of adequately trained personnel to limited laboratory facilities and intermittent pharmaceutical supply(4). Ethiopia, which has 123 million inhabitants, only has one radiotherapy machine, contrary to the International Atomic Energy Agency’s (IAEA) recommendation that a country have one radiotherapy machine per million people. Tikur Anbessa Specialized Hospital is the only tertiary hospital that delivers radiotherapy to all patients requiring it.

Studies suggest timely diagnosis of childhood cancer is basic in order to provide timely treatment before the disease prognoses and disseminate to other body parts, complicating the treatment needed and increase the time needed to heal(5). Accordingly, late presentation has been linked to an increased risk of death in children with cancer(5,6).

Studies imply, throughout their child’s diagnosis and rehabilitation, parents expressed confusion regarding their parenting techniques. They struggled with their new parental duties of nurturer and caregiver, unsure of how to treat and parent their child, and as a result, their parenting techniques were inconsistent(7–9). Other studies indicate parents suffer considerable level of guilt toward the child in the premise of causing the child’s disease through a variety of malpractices and situations after learning about the ethology of childhood cancer (10–12). Insufficient information about their child diagnosis, treatment and side effects(13), household economic hardship because of out of pocket treatment of the child, transport fee, loss of income from termination of employment and being unable to engage on other part time job (7,14,15), and healthcare system inadequacies(16) are considered as challenges that are faced by parents of child with childhood cancer. Majority of studies indicate psychological challenges while: anxiety, despair, helplessness, weariness, loneliness, having intense sentiments and reactions, as well as violence are the most regularly reported issues of parents or caregivers in research(15,17–20).

The main objective of the study was to look further into the lived experiences of Ethiopian mother with children suffering childhood cancer, from the mothers’ point of view. The study also aims to describe the challenges that mothers of a child with childhood cancer encountered in multiple dimensions, and document life style change made as a coping mechanism.

## Methods

The study was conducted in Tikur Anbesa hospital, the only tertiary hospital in Ethiopia providing radiotherapy services. Though the hospital has several expansion centers, there is still overcrowding owing to the enormous number of people coming from various regions of the country in need of the service. Patients and patient care attendants are hassled by the crowded surroundings. The other study setting is Tesfa Addis Parents Childhood Cancer organization, a non-profit organization covering all direct and indirect medical expenses incurred by children while they reside in the organization facilitated compound through the course of their treatment. The study was conducted from March - August, 2022 G.C.

For this research, a qualitative research approach was implemented. Interpretive phenomenology is the approach used. Interpretative phenomenological analysis is a method designed to understand people’s lived experience and how they make sense of it in the context of their personal and social worlds(21).Using a phenomenological perspective, it is attempted to comprehend the daily experiences of mothers parenting a child with Childhood Cancer.

The study procedure was rigorously followed throughout the course of the study. To enhance the evaluation process, all procedures have also been documented for confirmation and the assessment. Peer debriefing had been taking place while the data was being collected and analyzed. A continuous literature review was conducted along with the usage of different data sources, which included pictures, notes, and reflections from staff members of Tikur Anbesa Hospital as well as Tesfa Addis Parents Childhood Organization.

A snowball sampling was used to recruit mothers of children who have already been diagnosed with Childhood Cancer and now are receiving or waiting for treatment at Tikur Anbessa specialized hospital or who are temporarily residing at Tesfa Addis Parents Childhood Organization were the study participants. Mothers who are the primary caregiver for their biological child, who is medically diagnosed with childhood cancer, for at least two months after the initiation of treatment were eligible for the study. Mothers having different original residential area, varied educational status, and range of duration of treatment were recruited in order to grasp alternative perspectives. Staff at the centers assisted with the recursion process. Data collection was done through a semi-structured in-depth interview.

Both centers were visited multiple times before the data collection day in order to witness the macro-environmental factors study participants dwell in, to observe the services provided, to observe the interaction of study participants with their children, and to assess coordination. The prolonged stay of the researchers combined with the frequent visits allowed study participants to become accustomed to and gain confidence in the data collecting procedure.

The semi structured in-depth interview contained open-ended questions listed in the interview guide as well as probing questions. After confirming their willingness to cooperate and before audio recording the interview, a code was provided to each participant in order to keep the data confidential. The study location differed depending on the preference of participants and the conventionality of the space. The institution cafeteria was used for majority of the participants from Tikur Anbesa hospital as it has a good natural light to create a sense of ease to the semi structured in-depth interview. The cafeteria also had an open-air setup allowing the participants to talk in relaxed state of mind. As for the participants form Tesfa Addis Parents Childhood Organization the semi structured interview was conducted in one free room isolated from the administration office and other staffs as well as patients staying rooms contributing to the feeling of confidentiality to the study participants. The room had a quite atmosphere which contributed to the quality of audio recording and flow of thoughts.

Data saturation was regarded as the point at which there was no longer any possibility of obtaining new information, enough information to replicate the study had been gained, and further code generation was not viable. The duration of interviews ranged from eighteen minute to frothy three minutes.

Interpretative Phenomenological Analysis is used to analyze the data collected. In an effort to boost the data’s richness, notes from the interviews were added to the transcribed data during the transcription process. Since Amharic was the language of preference for the interviews, all transcriptions were done in Amharic language. Following transcription, translation was performed in order to get the data in the language of the study (English). All taped interviews, memos, and field notes are saved in a protected personal computer file. Analysis of data can be outlined in four steps

I. Multiple reading and making notes
II. Transforming notes in to emergent themes
III. Seeking relationships and clustering themes
IV. Writing up the study

The Addis Ababa University School of Public Health Research Ethics Committee, as well as the pediatric oncology department research ethics committee, provided ethical approval. Before performing every interview, verbal consent was taken after explaining the objective of the study and reading the consent. In order to ensure the privacy of participants, a code was used instead of their names, and data analysis was done in a way that would not expose the personal identification of participants.

## Result

From the participants in the study, ten mothers were married and two were divorced. None of the participants were disabled. All of the participants had no other children diagnosed with cancer. Four out of twelve mothers had only one child (the one who was diagnosed with childhood cancer), three mothers had two children, and the rest had three or more children.

**Table 1.**
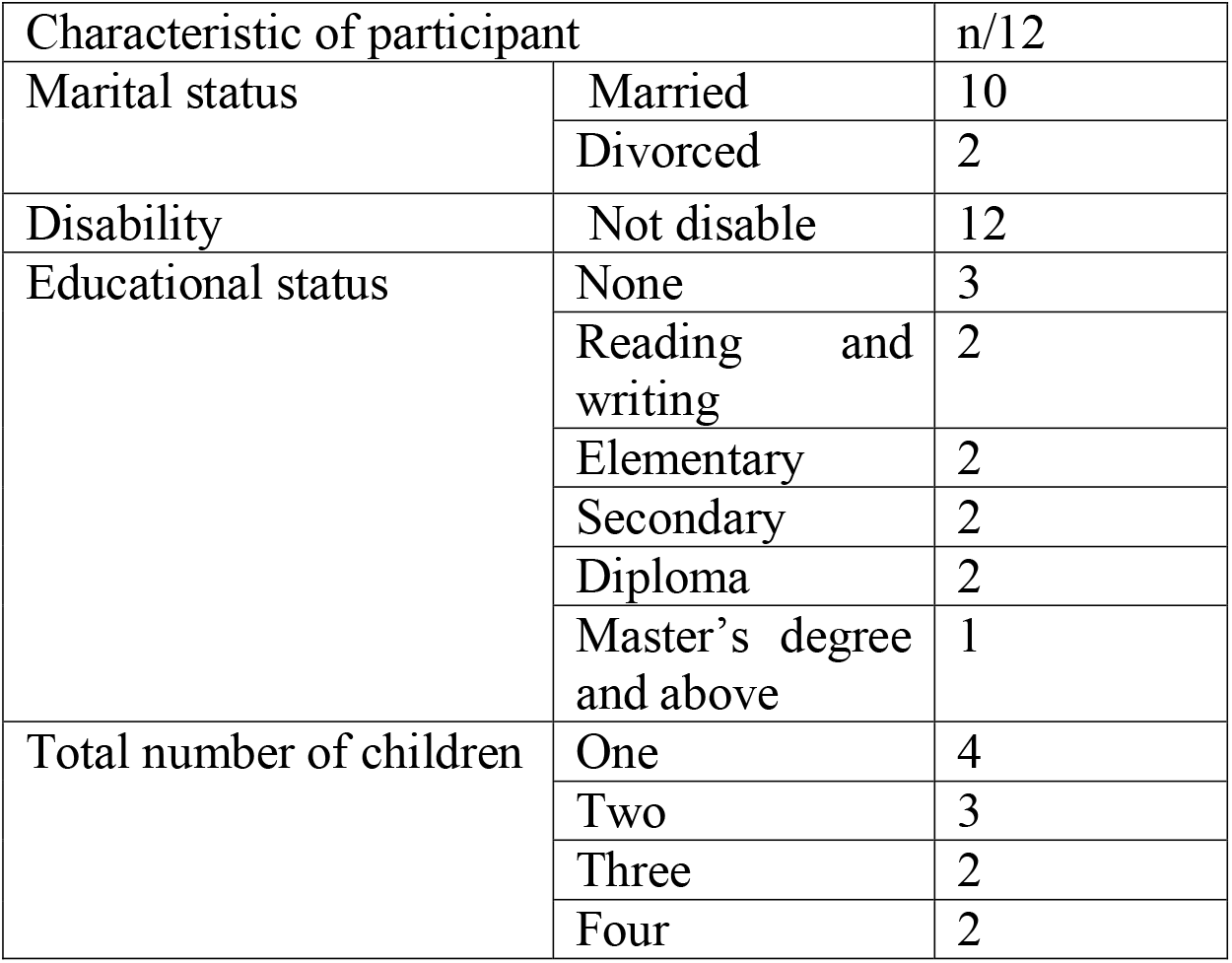
Socio Demographic Characteristics of the Study Participants.

According to the analysis adversity, coping mechanism and resilience are General theme that includes basic theme.

**Table 2.** Themes and sub-themes for The Lived Experience of Mothers of a Child with Cancer: in case of Tikur Anbessa Specialized Hospital and Tesfa Addis Parents Childhood Organization, Addis Ababa, Ethiopia.

| Theme | Sub-theme | Description |
| --- | --- | --- |
| **Adversity** | Caring related factors | encompasses challenging situations that arose while a participant nursed a child who had childhood cancer. |
|  | Family related factors | pertaining to any condition that weakens the family structure of participants.<br>For example: divorce, parenting withdrawal |
|  | Financial related factors | refers to any additional costs that participants have incurred as a result of their child's childhood cancer. |
|  | Information related factors | refers to a scarcity of adequate, reliable, and up-to-date information regarding childhood cancer |
|  | Emotional Issues | refers to the participant's emotions being adversely influenced by the child's childhood cancer diagnosis. |
|  | Problems related to spirituality | corresponds to skepticism and discontent with religion and God. |
| **Coping Mechanism** | Debt | refers to the total amount due to a financial institution or to any individual. |
|  | Sell house and properties | refers to exchanging money for owned properties to be able to cover medical expenses. |
|  | Social support | refers to verbal or nonverbal interactions that provide assistance |
|  | Emotional support | refers to the interaction leading study participants to believe that they are cared for, loved, and esteemed |
| **Resilience** | Adhere to treatment | denotes a determination to commit to the prescribed course of medical treatment in spite of adversity. |
|  | Anticipate to be an inspiration | corresponds with the participants' desire and hope to educate other mothers about the fundamentals of childhood cancer while also sharing their own story of making it through their child's treatment. |
|  | Feel expert | refers to the participants' notion that they are well-versed in childhood cancer as medical experts. |

### Theme 1 Adversity

This is one of the basic themes that emerged from the data analysis, which includes the difficulties mothers of children with cancer have, as well as the various hardship they confront throughout the medical situation. Based on the types of challenges theme is classified as problems related to caring, family, finance, information, emotion and spiritual.

### Caring related factors

Mothers of cancer patients confront difficulties in providing care while undergoing medical treatment. This section attempts to address the issues they face.

The limited number of facilities providing tertiary level care forces mothers to travel long way to Addis Ababa to get the treatment for cancer. This specific study includes study participants who originally reside in Gojam, Desse, Wolliso, Butajira, Alamata, Wollo, Gambella, Asela, Raya, Shinshicho, Ambo and Addis Ababa.

As attending their child’s treatment, mothers congregate with strangers; at such times, some of the children’s actions, and lengthy stay at the expense of other individuals or organizations, cause a sense of shame or awkwardness.

Also, another mother in Tesfa Addis Parents Childhood cancer organization said *“It is embarrassing to stay for so long on the expense of the organization*.*”(TAP02)*

Attending a child’s medical treatment necessitates being physically active for long periods of time, controlling the child not to touch any dirt material, constantly cleaning the environment, supervising the child’s activity, and so on. Furthermore, the numerous thoughts that run through a mother’s mind at this period cause mental fatigue. In addition, as part of care for the treatment mothers provide physical support for their child; which might include holding the child in a position favorable for him/ her, carrying the child on the back to take him/ her to different spots, carrying them to sleep, help them to use the rest room and so on. Providing a physical support for extended period of time asks a strong physical coordination. Though it is tiering mothers have a difficult time trusting people to care for their child. They believe that others are hesitant because of the child’s illness. This will urge them to undertake all of the caring on their own. Furthermore, mothers exert an overwhelming amount of effort to make their sick child’s siblings and other family members’ surroundings better. They will accomplish this by providing comforting conversations and positive energy when they are most in need of it. Meanwhile mothers face resentment from their spouses, family and hospital staff members.

*“At the time, my daughter couldn’t sit still, so I would spend hours bending and maintaining her in a comfortable position, I have been having pain whenever I try to bend down since then*.*”* said a mother.(*TAP01*)

A mother from remote side of Ethiopia said that *“And after he takes the medicine, he refuses to eat, has a fluctuating high temperature, then recovers. He would still not eat when we returned home from the hospital. And when I force feed him, he vomits blood even after the food is gone, so I must feed him again*.*”(TAP03)*

Mother who have been attending the treatment of her child for four years said *“I have a hard time to trust on fetching her medication without my supervision*.*”*

A single mother whose child’s father is a health professional said, *“Her father helped me until I came to Addis Ababa, but after that, he told me that she would not be saved and would not have much support*.*” Once I asked her father to send me 500 birr every month for her, but he literally said, “I will not send you 500 birr for a dyeing child; I will use it for myself. (TAP04)*

### Family related factors

The treatment of the child seems to create a topic for some families to fight over. Not understanding the situation, they are suffering alone in their solitary confinement, some will consider their wife having an affair with another man in the city. Not only that, but the physical distance created by the marriage caused husbands to seek out another wife figure in their home and replace the mother who is attending their child’s treatment with another woman within their reach.

A mother who stayed long in Tesfa Addis parents childhood organization due to the unstable political condition said *“I left my house. When I stayed here, he thought I was sitting in comfort. He thinks that the abuse I am going through here is a convenience, while I am here to care for our child, he has married another one there*.*”(TAP03)*

Mothers stated that not every family has a single child; some have two or more. When caring for a child with cancer, mothers are compelled to emphasize the sick child and reduce their maternal obligation for their other child/children.

A mother of four and a new mom at the time of interview said*” I even leave my other children with my weak mother. I don’t even have someone to take my children to and from school*.*”(TA02)*

### Financial related factors

The inability to make your payments on time or fulfill your basic demands is a sign that you have a financial problem. The addition of costs for a child’s cancer treatment has the potential to upset one’s equilibrium between income and expense.

Currently, Ethiopian government facilitates citizens accessing medical care through the use of health insurance, though the insurance card makes most of the services provided in government health centers free, there are some services that are either temporarily or permanently unavailable in government health facilities. As a result, mothers would have to pay for the services out of their pocket.

A mother who has no medical insurance before said*” When I first came, his family and mine gave me what they had. I came here and we didn’t have health insurance then, so all that money was spent on tests, and we didn’t have any left over for the treatment. When we arrived, we were wondering what to do with the money we had saved from our son’s treatment; But we went to six hospitals for this child’s examination and run out of money before the treatment even start*.*”(TAP06)*

Additionally, participants are also exposed to expenses that are not directly connected to receiving medical care, such as lodging expenses for medical care, personal care costs, and travel costs.

*“There is a choice of food, you should be careful about his hygiene because now his resistance to disease is extremely poor, so he needs a lot of care, and today, as you know, everything is expensive, it costs a lot,”* said a mother from Addis Ababa. (TA05)

### Information related factors

Most mothers are unaware about cancer, its treatment, necessary precautions, the severity of the sickness, and the length of therapy. As a result, mothers begin their child’s treatment without adequate preparation.

Mother, who had no prior exposure to cancer, said, *“When I came here, I thought that her eyesight would never be good (she would not see again). They would just throw her eyeball out and send me back to my home soon, but this is different. (TA04)*

Moreover, unreliable information about cancer is spread, which discourages mothers from following the treatment and simply waiting for their child to die.

Mother who had a divorce said *“my father suggested to quite trying. He said “I have been asking so many people about it here, none of them survived and their families just wasted their fortune on them for nothing”. So I hesitated to come back to Addis Ababa*.*”(TAP04)*

### Emotional Issues

Given the severity of cancer, mothers experience anxiety, helplessness, impatience, forgetfulness and regret.

Participants not only does the severity of the sickness make them anxious, but so does their exposure to new surroundings in the metropolis.

A mother with prior harsh life experiences said, *“Cancer is confusing*.*” “Even on this campus, many children who we said were safe have died, and I am afraid that my child will die suddenly when I say he is safe*.*”(TAP05)*

When mothers notice the duration, it takes for the entire medical treatment, the emotional load, and other treatment needs, they become impatient.

A mother who expressed she was losing her patience said, *“In general, the disease itself is challenging; the disease is severe; the disease tests patience*.*” (TAP05)*

While the mind is processing a lot of thoughts at once, the capacity of memory drops. As mothers who care for their children with cancer face a lot of new information and emotion, they tend to forget some of it.

An emotional mother said crying *“I am starting to forget what is being said to me. If we were there in the countryside, my son would be dead and forgotten by now. Even after the operation, my son could not recover quickly and stayed in the ICU for three days. I don’t remember anything other than that*.*”(TAP06)*

When evaluating their child’s current circumstances, mothers grieve the things they did and did not do in the past to improve their child’s health outcome.

A mother who waited for five months in different government hospitals waiting for treatment said *“Now, when I think about it, I think she might have the chance to be healed if she had gotten treatment as soon as we arrived*.*”(TA04)*

### Problems related to Spirituality

Mothers’ express dissatisfaction or annoyance about the child condition and blame God for it. Mothers frequently mention that they are punished with their child’s condition. They even question why would God allow such a suffering in to their lives.

After watching her daughter suffer, her mother said, *“I used to say, “God, why are you torturing her?” “She is better off dead*.*”(TA01)*

### Theme 2: Coping mechanisms

#### I. Debt

Mothers owe money for the child medical follow-ups and for some expenses that come along with the medical care. The debt could be from family, friends or institutions that offer financial debt.

*“But my husband suggested that your parents should not sell cattle, therefore I will borrow 25,000 birr from the cooperative organization,” a low-income mother replied*.*(TAP03)*

#### II. Sell house and properties

Children are the priority of mothers that cannot be won with money, thus in such circumstances, they sell their home as well as their asset to fund medical expenditures.

*“When we started treatment, we sold our house in the city, but the money ran out immediately. My daughter would have died because we had no other assets left*.*” Stated the mother (TA01)*

*“We have no money in the bank, our money is our sheep and cattle; We sold our cattle and started attending the treatment because we did not have enough money. Our child was our priority that we cannot get by money*.*”* said a mother from rural area.*(TAP01)*

#### III. Social support

Apart from the financial support, participants also get social support from family members, friends, neighbors, staff of hospitals, and strangers. Having such support helps mothers gain hope and keep seeking medical as well as spiritual solutions.

□*May God bless the medical staff with a kind heart, she made a lot of sacrifices with me. With the help of the doctors and her, my son has reached the level he is now*.□ *Said a grateful mother. (TAP06)*

#### IV. Emotional Support

Getting emotional support is key to resolving the anxiety, helplessness, impatience, regret, and many other emotional stresses caused by children’s medical conditions. The emotional support can be any kind and can be fetched from family, friends, health professionals following their child’s treatment, and even sometimes strangers.

The mother, who has been attending her only child’s treatment, said, *“What I think has helped me is having my husband with me*.*” He tells me not to be afraid; we will pray together. My mother is the one who handles my child when I have to leave the house. Sometimes our families come together and we play; in that case, it will be a little easier for us. “I miss the days we spend together with the family, especially since there is no work on Saturdays and Sundays*.*”(TA05)*

### Theme 3: Resilience

The multiple types of assistance that mothers receive from various sources assist them in gaining strength and moving forward. Even when mothers have a difficult moment, they keep fighting and pushing forward.

#### I. Adhere to treatment

Without hope of a cure, it is hard to continue the treatment of a child. As the treatment progresses, they become more determined to complete it.

A mother who can be a model for treatment adherence said, *“Everyone says that I shall stop caring for her because it is only a matter of time before she dies*.*” But in the middle, God told me not to give up hope and to continue the medicine. Now my daughter has started school*.*”(TA01)*

#### II. Anticipate to be an inspiration

Hearing the testimony of other mothers who have had their children recover motivates them to share their experience in the future for the benefit of others.

*“I did not lose faith in all of this because I imagined she would grow up tomorrow and benefit me and the government and subsequently become an example for others,”* said the mother of a daughter with leukemia. (TAP04)

#### III. Feel expert

Mothers develop basic knowledge about their child’s medical condition. As they get familiar with the medical terms used by the health professionals and the steps of the treatment, they start to feel like they are medical professionals themselves.

Mother who withdrawn her education from grade eight said*” I can now say I am a doctor myself and for my daughter*.*”(TAP04)*

## Discussion

The experience of parents who have a child with cancer was examined in this study. Mothers had the opportunity to share their experiences from their initial encounters when they witnessed their child presenting symptoms of a disease until they received a diagnosis of childhood cancer and received guidance on how to proceed with treatment. Their understanding of childhood cancer and its effects on their social, psychological, and spiritual wellness, as well as their coping methods and unmet needs, the study’s findings highlighted both the negative and positive aspects of parenting a child with cancer. Positive experiences were also discovered, which is parallel to another study on childhood cancer, despite the fact that it largely concentrated on the challenging parts of the experience(10,17).

This study illustrates the difficulties mothers encounter when caring for their child who has cancer in childhood. The study’s participants noted that their extended hospital stays created a communication gap which sometimes led to divorce. Mothers felt obliged to abandon parenting their other children in order to focus on caring for their child who was receiving treatment for childhood cancer(15–17).

The most prevalent problem category observed in both this study and related studies is financial problems. Even though Ethiopia is developing a medical insurance system, the coverage is not adequate. There is also an extended list of indirect medical expenses, which at times may be more than the direct medical expense(7,22–24).

Participants in the study who are from highly remote regions of Ethiopia indicated that before they arrived at Tikur Anbesa for treatment, they knew very little to no information about childhood cancer. Because they receive their information about cancer and its treatment informally from unreliable information sources, the majority of mothers have an inaccurate understanding of the disease and its treatment, as well as a very tense and depressing mindset(12,17).

The majority of study participants affirmed their profound sorrow for the situation they are in and their high level of anxiety over losing their irreplaceable son or daughter. Mothers in the study also expressed regret about the time they wasted waiting to begin their child’s treatment. The study additionally explores the supplementary psychological problems of despair and helplessness. Mothers begin to lose patience with following the treatment as the child’s health begins to deteriorate(10,20,25,26).

Despite having diverse religions, the study participants all beseech to God to explain why this is happening to them. Mothers plead God for mercy because of the enduring notion that all illnesses are retribution for the sins they do. The results of comparable studies can also be compared to such(22).

Also covered in the study is resilience in the face of challenges. The mothers manage to have the fortitude to stand firm and conquer the difficulties for their child who has childhood cancer even though they endure a variety of problems each day. One method to demonstrate their resiliency is by adhering to their child’s medical treatment(18).

## Conclusion

The study provides crucial insights into the experiences of mothers of children with childhood cancer. The diagnosis of childhood cancer has a profound impact on mothers, and it elicits various reactions from them. Investigate the emotional, mental, and social drivers of caregiving, emphasizing important factors such as financial pressure, insufficient quality medical treatment, emotional disturbances, and strategies of coping with their children by the mothers.

Health facilities that can provide advanced medical care should be built in every region of the country to increase the accessibility of quality medical care. Medical professionals should undergo recurrent on-the-job training on screening pediatric cancer and primary care to minimize the misdiagnosis of childhood cancer at any level of health care center. Additionally, establish counseling and support schemes for mothers to cope with cancer therapy for their child. Organize community awareness campaigns to increase understanding, reduce stigma, and promote early treatment.

## Data Availability

All data produced in the present study are available upon reasonable request to the authors

## Notes

### Competing Interest Statement

The authors have declared no competing interest.

### Funding Statement

Addis Ababa University Female Scholarship program

### Author Declarations

Research Ethics committee of Addis Ababa University School of Public Health and Research Ethics Committee of Addis Ababa University pediatric oncology department waived ethical approval for this work

